# Upsurge in Hospitalization of Pediatric Patients with Severe Acute Respiratory Infections in Kolkata and Surrounding Districts Caused by Recombinant Human Respiratory Adenovirus Type B 7/3

**DOI:** 10.1101/2023.04.10.23288279

**Authors:** Agniva Majumdar, Ritubrita Saha, Ananya Chatterjee, Rudrak Gupta, Ratul Datta Chaudhuri, Alok Kumar Chakrabarti, Mamta Chawla-Sarkar, Shanta Dutta

## Abstract

Globally, different genotypes of human adenoviruses are associated with outbreaks of acute respiratory infection though such evidence is lacking from India. In the present study, we report a sudden increase in the positivity of respiratory adenovirus among hospitalized children with acute respiratory infection from Kolkata and the surrounding districts of West Bengal, India, from December 2022 till date. A sharp rise in the positivity rate of respiratory adenovirus was found which ranged from 22.1% in early December 2022 to 52.6% in mid-March 2023. The overall positivity was 40.4% during the period and children in the 2 to <5 years (51.0%) age group were mostly affected. Single infection with adenovirus was found in 72.4% of cases while co-infection with Rhinovirus was the maximum (9.4%). Around 97.5% of positive cases required hospitalization. Cough, breathlessness, and wheeze were the most common clinical features among positive patients. Phylogenetic analysis of the hexon and fiber gene of all the sequenced strains revealed HAdV-B 7/3 recombination with more than 99% homology within themselves. This report of a respiratory adenovirus outbreak in West Bengal, India causing severe illness in the pediatric population underscores the need for regular monitoring of the circulating strains.

## 1 Introduction

Human adenoviruses (HAdV) are non-enveloped icosahedral viruses with double-stranded DNA which belong to the genus *Mastadenovirus* and family *Adenoviridae*. There are more than 90 serotypes of HAdV classified into seven species, designated A to G, based on their serological and molecular characteristics^1^. HAdVs cause a wide variety of diseases across all age groups - responsible for 5-10% of pediatric and 1-7% of adult infections^2^. Species D (8, 19, 37) usually causes conjunctivitis; species F (40, 41) is associated with gastroenteritis; and species B (3, 7, 11, 14, 16, 21, 55), C (1, 2, 5, 6), and E (4) cause respiratory diseases^3^. Infection severity is usually associated with different HAdV serotypes.

HAdVs causing acute respiratory infection (ARI) are mild to moderate in most cases, whereas the cases of fatalities due to HAdV pneumonia were reported in previously healthy children and adults – particularly among the immunocompromised with fatality rates up to 50%^4^. Respiratory HAdVs are highly contagious as the transmission occurs through respiratory secretions and person-to-person contact – increasing its outbreak potential^5^. The prevalence of HAdVs in Asia with ARI has ranged from 0.8 to 11.30%^3,6^. Several outbreaks of HAdV have occurred in China, Korea, the US military, Singapore, and Malaysia^5,7–9^. Several other countries like Japan, Canada, Paraguay, Argentina, Turkey, the UK, Brazil, Kuwait, Cameroon, Thailand, and Colombia have also reported severe HAdV infections^10,11^. Species B HAdVs 3, 4, 7, 14, and 55 are mainly associated with these severe respiratory disease outbreaks^1^. In India, there are limited reports on HAdV although circulation of 3, 7, and 2 types was reported in Eastern and Southern India^12,13^. However, there is no documented report of an outbreak of severe acute respiratory infection caused by HAdV.

The Regional Virus Research and Diagnostic Laboratory (VRDL) at ICMR – National Institute of Cholera and Enteric Diseases (NICED), Kolkata, is a regional laboratory of the VRDL network under DHR-ICMR involved with timely identification, reporting, and research on viral diseases and other agents of public health importance and epidemic potential. As part of routine screening of respiratory pathogens, VRDL-NICED witnessed a sudden increase in HAdV positivity among the hospitalized cases of ARI from December 2022 onwards which by mid-March 2023 had increased to 52.6%. In the present study, we report the clinical findings of the patients infected with HAdV and the character of the strain causing the infection.

## 2 Materials and Methods

### 2.1 Source and collection of samples

As per WHO case definitions, ILI is an ARI with measured fever of ≥ 38°C and cough with onset within last 10 days; SARI is an ARI with history of fever or measured fever of ≥ 38°C and cough, with onset within the last 10 days and requires hospitalization. Nasopharyngeal and/or oropharyngeal swabs collected in Viral Transport Medium from patients with ILI/SARI were received at VRDL-NICED from different hospitals of Kolkata and the adjacent districts of West Bengal maintaining the cold chain – mostly tertiary care hospitals of Kolkata like Dr. B.C. Roy Post Graduate Institute of Paediatric Sciences, Medical College & Hospital, Ramakrishna Mission Seva Pratisthan, Calcutta National Medical College & Hospital, and Institute of Post Graduate Medical Education & Research. The vials containing the sample were vortexed, followed by centrifugation at 1500 rpm for 5 min before proceeding with nucleic acid extraction.

### 2.2 Detection of HAdV

Nucleic acid extraction was performed using ThermoFisher MagMAX Viral/Pathogen Nucleic Acid Isolation Kit followed by detection of the panel of respiratory viruses (Influenza A H1N1, Influenza A H3N2, Influenza B, SARS-CoV-2, Respiratory Syncytial Virus, Adenovirus, Rhinovirus, Parainfluenza virus, human Metapneumovirus) by real-time reverse transcriptase-PCR (ABI 7500, Applied Biosystems, USA). The adenovirus was detected by a pre-published primer-probe set and was considered positive if the reaction growth curve crossed the threshold line within 35 cycles^14^.

### 2.3 Genotyping of HAdV

Samples screened positive for HAdV were further used for genotyping. A total of 40 samples were genotyped: 10 samples each month from December 2022 to March 2023. Hypervariable regions 1-6 of the hexon gene and tail-knob region of the fiber gene were sequenced for HAdV genotyping and phylogenetic analysis of circulating adenovirus strains. Three sets of primers targeting the hexon gene and fiber gene were used for the amplification of the adenovirus genome (**Supplementary Table 1**). HAdV-specific PCR products were purified using Qiagen PCR purification kit. Sequencing was performed) in ABI Prism 3730 Genetic Analyzer (PE Applied Biosystems, Foster City, CA, USA) using ABI Prism Big Dye Terminator Cycle Sequencing Ready Reaction Kit v3.1 (Applied Biosystems. All 40 sequences were analyzed through NCBI-BLAST (National Centre for Biotechnology Information - Basic Local Alignment Search Tool) server on the GenBank database (release 143.0) to determine the genotypes.

### 2.4 Phylogenetic Analysis

Phylogenetic analysis was performed on all the hexon and fiber sequences. The gene sequences were submitted to the GenBank database under the accession number OQ686796-OQ686799, OQ709803-OQ709858, and OQ709862-OQ709881. Amplified hexon and fiber sequences were aligned with other HAdV sequences available in NCBI GenBank using the MUSCLE method of MEGA (Molecular Evolutionary Genetics Analysis) software (version 11). Phylogenetic trees were constructed using the MEGA software and the substitution model was based on the maximum likelihood statistical method that produce the lowest BIC (Bayesian information criterion) and AICC (Akaike information criterion (corrected)) scores through the model testing parameter tool. Models used for phylogenetic tree construction were GTR+G+I and GTR+G for hexon and fiber tree respectively (1000 bootstrap replicates). Homology between the study strains and other reference strains was assessed using pairwise sequence alignment in LALIGN software (EMBnet).

### 2.5 Data analysis

Demographic and clinical details were collected from the accompanying test request forms and ILI/SARI information sheets and analyzed by time, place, person, and clinical features. Statistical analysis was done using Epi Info version 7.2.5.0.

## 3 Results

The weekly distribution of cases from December 2022 to March 2023 as depicted in **Figure 1** shows the positivity rate increased from 22.1% in early December 2022 to 52.6% by mid-March 2023. A total of 3115 cases of ARI were screened for HAdV from 04 December 2022 to 18 March 2023, out of which 1257 (40.4%) tested positive. Most of the HAdV-positive cases (n=1154; 91.8%) were from Kolkata and the surrounding districts of Howrah, Hooghly, Nadia, North, and South 24 Parganas. Positivity in males was 41.4% (n=783/1891) and that in females was 38.7% (n=474/1224). The age of the HAdV-positive cases ranged from 1 month and 2 days to 78 years, with a median of 1 year and 7 days. The odds of HAdV infection in cases <10 years of age was 4.46 times than those aged 10 years or more (odds ratio 4.46; *P* value <0.0001) with the highest positivity in the 2 to <5 years (51.0%) age group (**Table 1**). Out of the 1257 positive cases, 910 (72.4%) cases had only HAdV infection while 347

**Figure 1:**
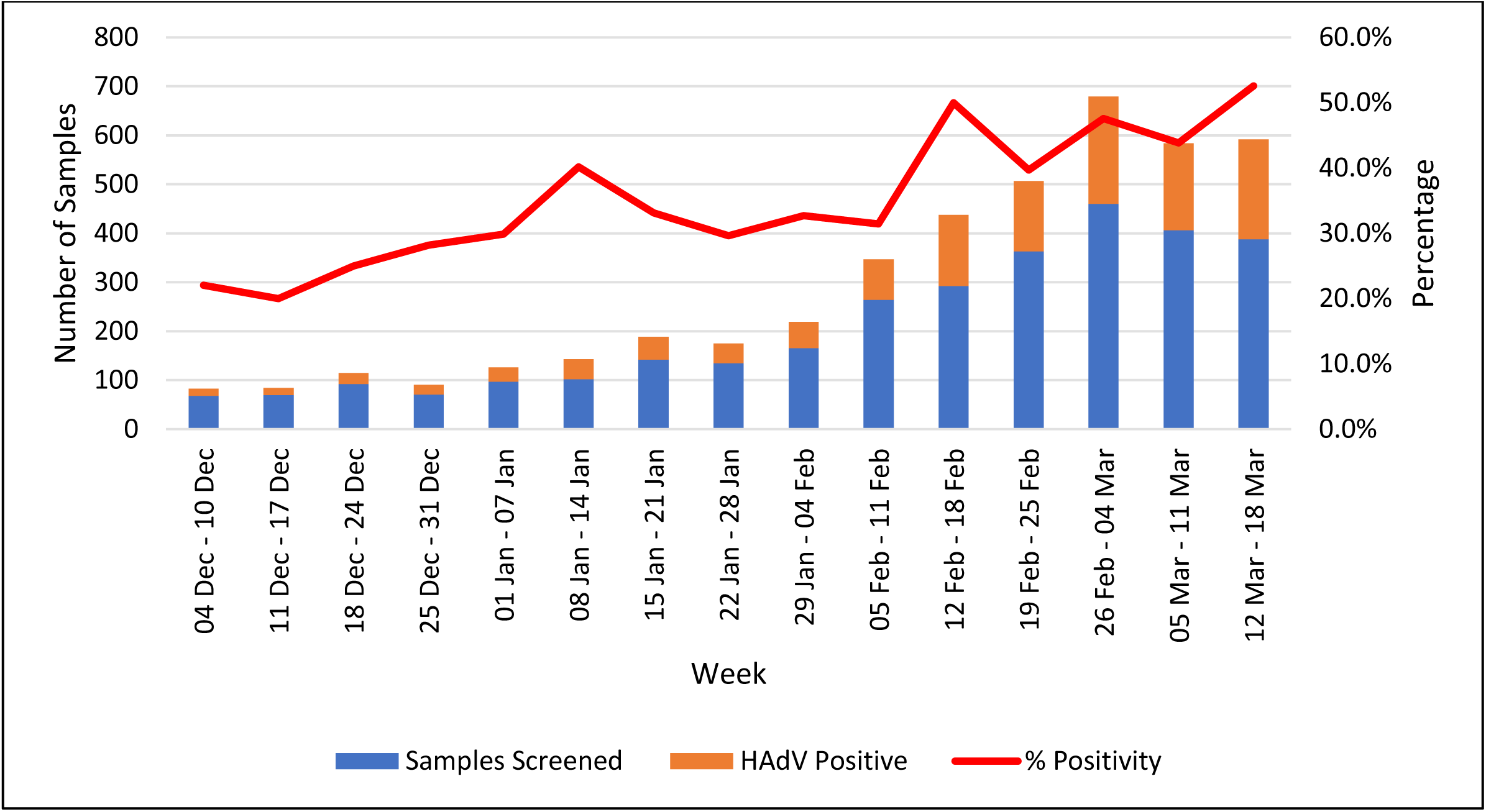
Temporal distribution of human respiratory adenovirus-positive cases from Nov 2022 to March 2023

**Table 1:**
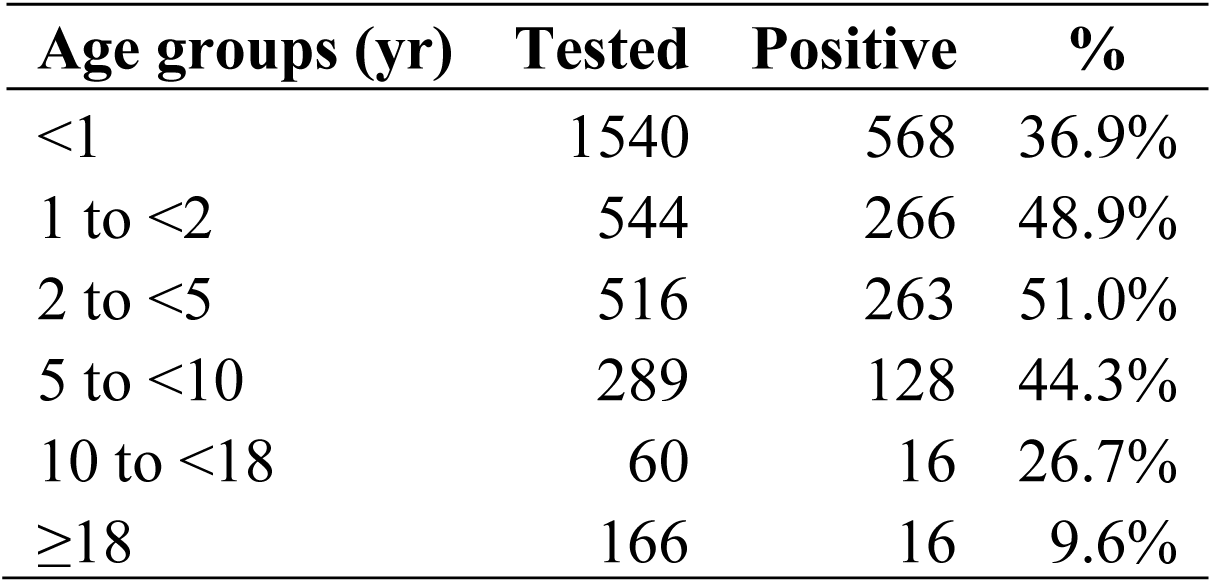
Age distribution of the HAdV-positive cases (n=1257)

(27.6%) cases had co-infection with other respiratory viruses. The most frequently co-infected virus is Rhinovirus (n=118; 9.4%), followed by Parainfluenza virus (n=87; 6.9%), Influenza A H1N1 (n=66; 5.3%), human Metapneumovirus (n=36; 2.9%), Respiratory Syncytial Virus (n=21; 1.7%), Influenza B (n=16; 1.3%), and Influenza A H3N2 (n= 3; 0.2%). Out of all the positive cases, 97.5% (n=1226) required hospitalization. **Table 2** depicts the associated clinical features: cough (83.1%) was the most common symptom followed by breathlessness (62.4%), nasal discharge/stuffiness (51.3%), sputum production (26.6%), vomiting/nausea (17.9%), seizures (6.1%), earache/discharge (3.3%), and hemoptysis (2.2%). On examination, wheeze (75.4%) was the most common sign followed by crepitations (52.4%), lower chest in-drawing (45.4%), nasal flaring (43.6%), accessory muscles used in breathing (34.1%), grunting (13.9%), stridor in calm patients (9.2%), and apnea (6.1%).

**Table 2:**
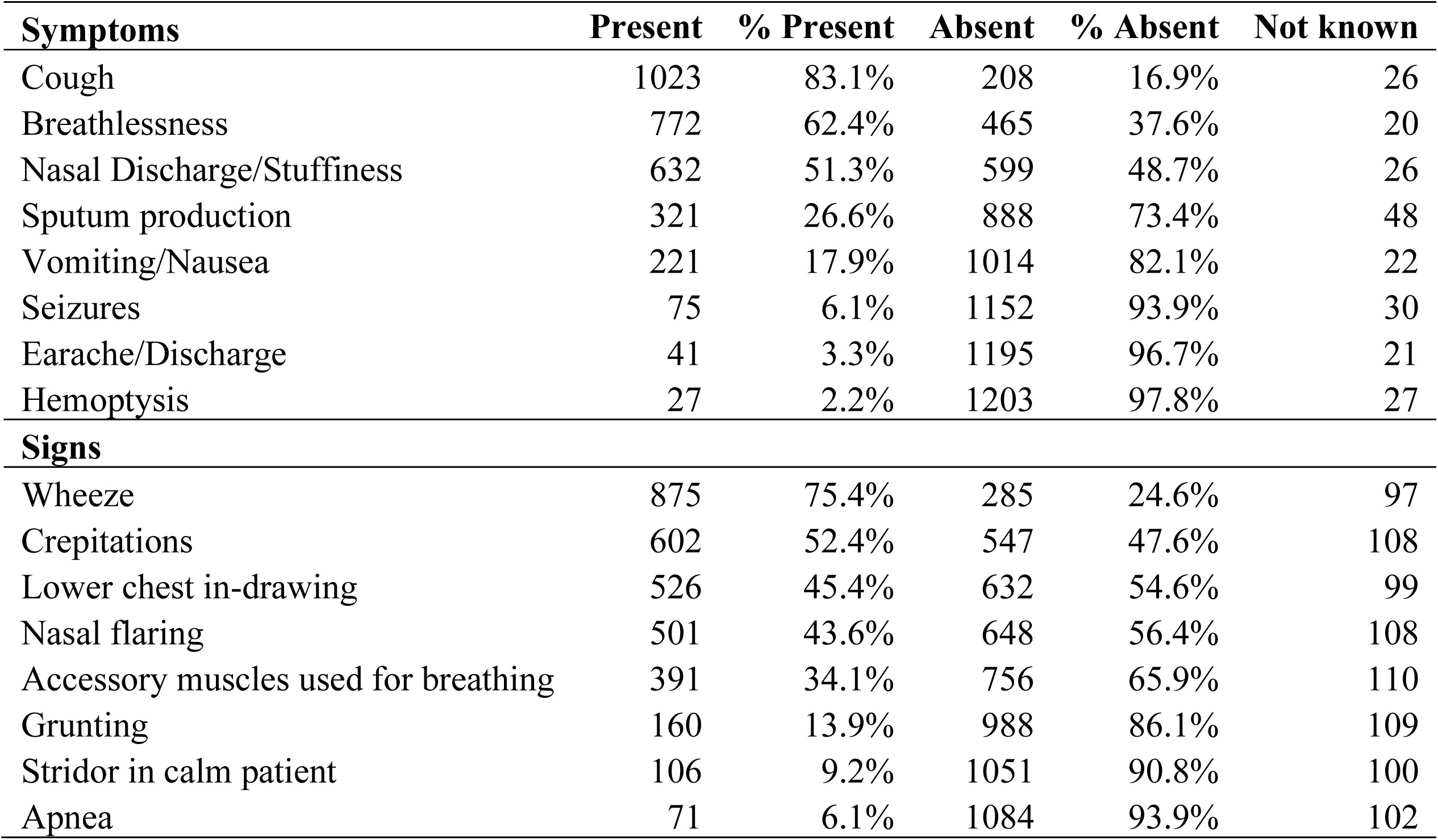
Clinical features associated with the HAdV-positive cases (n=1257)

The 40 samples were genotyped and characterized by phylogenetic analysis for the hexon and fiber genes. Hexon gene of the 40 representative strains clustered with the HAdV type B7 strain. These strains also shared ∼99.8-100% similarity with the HAdV-B 7/3 type recombinant reference sequence from Argentina (JN860676.1) and ∼98-100% similarity with UK, China, USA, and the two vaccine strains, (AY495969.1 and AY594256.1). The hexon gene of these 40 HAdV strains was closely related to the type 7 prototype strain Gomen (∼95%) and distant to the type 3 prototype strain GB (∼88.5%) (**Figure 2A**).

**Figure 2:**
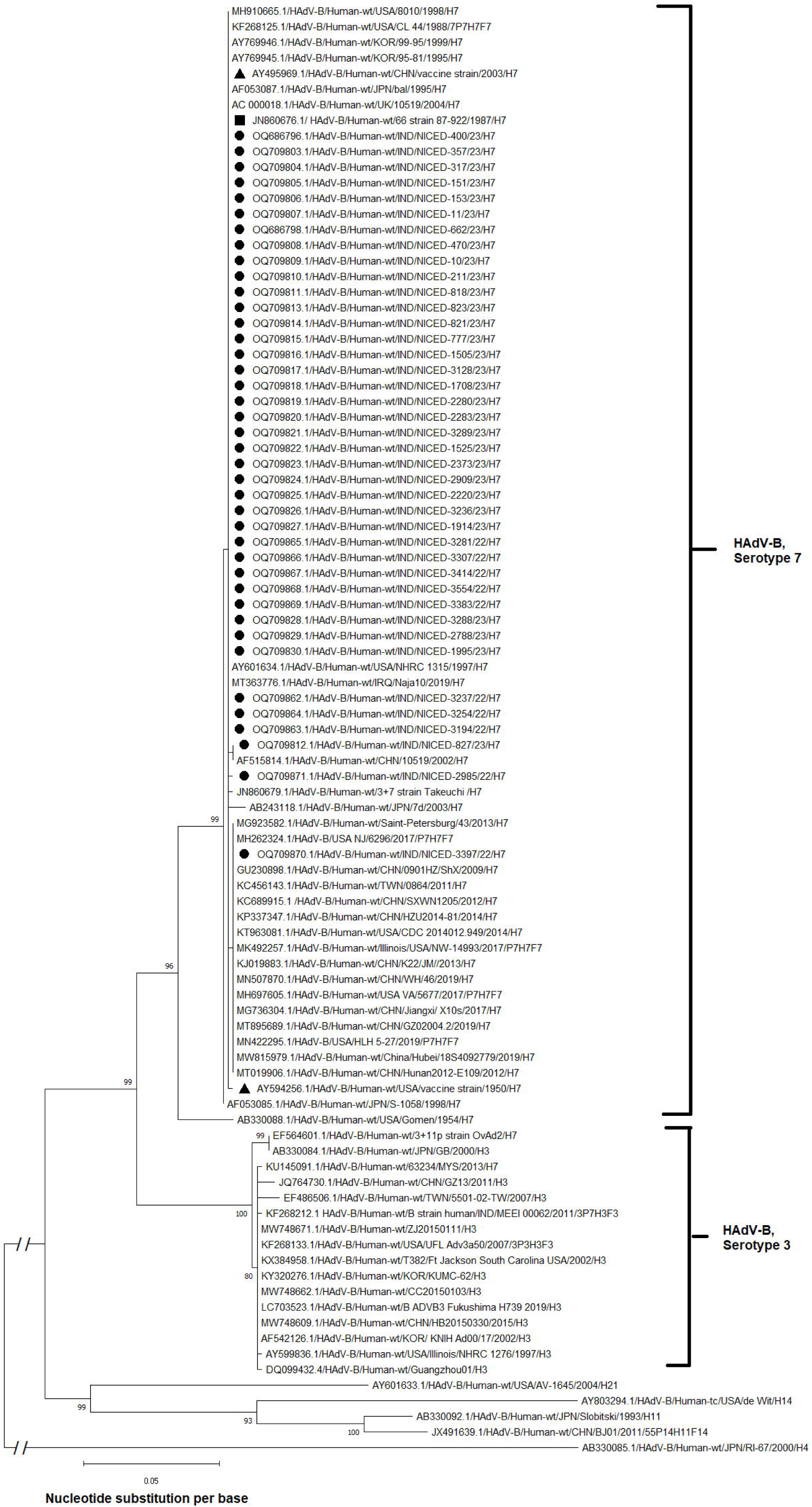
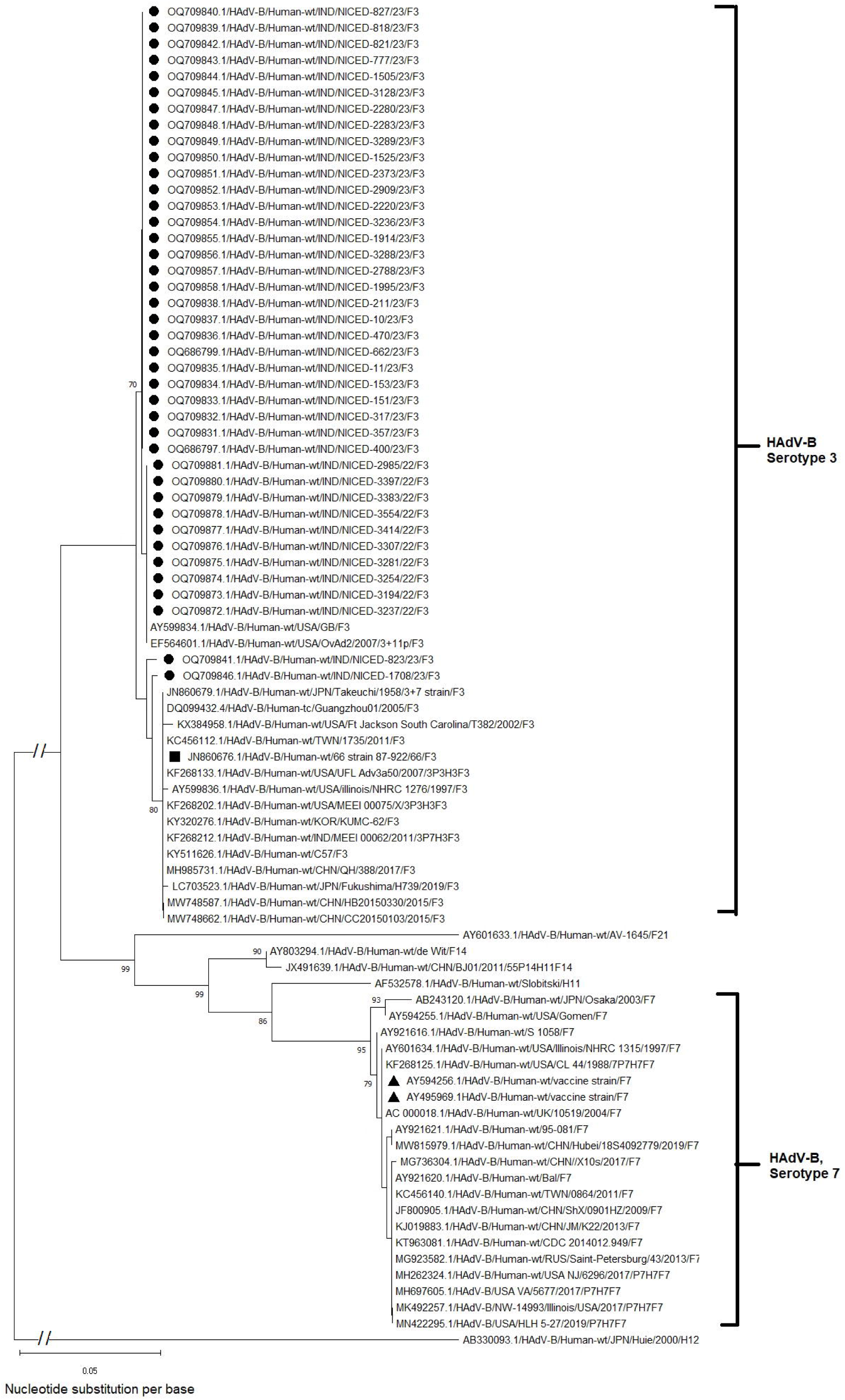
A, Phylogenetic dendrogram based on nucleotide sequences of Hexon gene of Human Adenovirus isolated during the outbreak in Dec 2022 – Mar 2023, with other known strains of different serotypes. The representative strains have been marked with a solid circle (●). The HAdV-B 7/3 recombinant reference strain has been shown with a solid square Scale-bar (■), and Vaccine strains have been marked with a triangle (▲). Scale-bar, 0.05 nucleotide substitutions per site. Bootstrap values of less than 70% are not shown. B, Phylogenetic dendrogram based on nucleotide sequences of Fiber gene of Human Adenovirus isolated during the outbreak in Dec 2022 – Mar 2023, with other known strains of different serotypes. The representative strains have been marked with a solid circle (●). The HAdV-B 7/3 recombinant reference strain has been shown with a solid square Scale-bar (■), and Vaccine strains have been marked with a triangle (▲). Scale-bar, 0.05 nucleotide substitutions per site. Bootstrap values of less than 70% are not shown.

All 40 samples sequenced for fiber gene clustered with the type B3 reference sequences. The fiber gene of the study strains was ∼98.9-99.4% similar to the recombinant strain (HAdV-B 7/3) and strains from Japan and USA (∼98.3-99.3%). All the fiber sequences reported in this study showed ∼99.1-99.8% similarity with type 3 prototype strain GB and they were found to be distantly related to the type 7 prototype strain, Gomen (∼63.7%). Results confirmed that all 40 strains are recombinant containing serotype 7 hexon protein and serotype 3 fiber protein (**Figure 2B**).

## 4 Discussion

HAdV-associated respiratory disease outbreaks are frequently reported across the globe. China alone had witnessed several outbreaks of Adenovirus^7,8^. Outbreaks in military and air force training centers were also reported in Korea and Texas, USA^9,15^. This study reports an outbreak of human adenovirus infection among children seeking hospital care in Kolkata and the surrounding districts of West Bengal, India. The rise in HAdV-positive cases was observed from December 2022 onwards with peak positivity in mid-March 2023 at the time of this report. The average positivity in the previous months from January – November 2022 was 4.8% (data not shown). HAdV infections mainly occur in children younger than 5 years. In this study, the maximum number of infected patients was also in this age group. HAdV accounts for 4%-10% of cases of pneumonia, 2%-10% of cases of bronchiolitis, and 3%-9% of cases of croup with around 20% of children and new-born suffering from pneumonia^16^. In this report, 97.5% of cases had severe clinical presentations and were admitted to the hospitals. The official report from the West Bengal Government stated only 19 deaths due to HAdV out of 13,061 admitted ARI cases from January to mid-March 2023^17^. But there may be gross underestimation^18^.

Generally, outbreaks of respiratory HAdVs were associated with species B, C, and E. Previous reports suggested that among the HAdV-B serotypes, 7 and 3 were the most potent cause of an outbreak^15,16^. They were associated with increased rates of hospitalizations although HAdV-B type 7 was more severe^1^. Recombination in adenovirus frequently occurs between strains of the same species rather than interspecific recombination. This study detected HAdV-B 7/3 recombinant virus in all the genotyped patient samples across the 4 months. These strains showed >99% homology within themselves suggesting it to be the outbreak strain probably from a single source. To our knowledge, an outbreak of HAdV with such a recombinant strain has not been reported in West Bengal, India. The HAdV-7h strain 87-922 which was isolated from a fatal case of pediatric pneumonia in Argentina in 1987 was a similar recombinant strain with a type 7-like hexon and a type 3-like fiber^19^. Portugal also reported an outbreak in 2004 associated with a similar 7/3 recombinant strain which was highly virulent and transmissible^20^.

Overall, in the study human adenovirus recombinant strain HAdV-B 7/3 has been implicated as the causative agent of the present outbreak of acute respiratory infection among children less than 5 years seeking hospital care in West Bengal. Our report emphasizes the need for in-depth studies on the impact of recombinant variants in the pathogenesis of human adenoviruses and the outcome of the patients.

## Data Availability

The data that support the findings of this study are available in GenBank at https://www.ncbi.nlm.nih.gov/genbank/ under the Accession number OQ686796-OQ686799, OQ709803-OQ709858, and OQ709862-OQ709881. The clinical data not mentioned already in the manuscript that supports the findings of this study is available from the corresponding author upon reasonable request.

## Author contributions

AM coordinated the collection of samples from hospitals and analyzed the clinical data. RS did the sequencing and phylogenetic analysis. AC and RG processed and screened the clinical samples for the presence of respiratory viral pathogens. RDC performed the amplification of PCR products and sequencing. MC, AKC, and SD conceptualized, designed, and monitored the study. AM and RS wrote the draft. MC and SD critically reviewed and edited the manuscript. All authors have read and agreed to the published version of the manuscript.

## Acknowledgment

The authors acknowledge the Department of Health Research (DHR), Indian Council of Medical Research (ICMR), ICMR-National Institute of Virology, and the Government of West Bengal for providing financial and logistic support for this study. RS was supported by junior research fellowship from University Grants Commission (UGC). The scientific and technical staff working at VRDL-NICED (Ashis Debnath, Pradip Kumar Jana, Soumen Mukherjee, Akash Ghosh, Sutapa Hazra, Bidisha Das, Chinmoy Mondal, Satyabrata Ghorai, Souvik Kar, Avisek Sinha, Amit Kumar Dutta, Asish Kumar Jana, Kartick Chandra Mondal, Biswajit Dey, Nayan Basuli and Bithin Banerjee) are acknowledged for their contribution in processing, testing, and reporting of the clinical samples.

## Data availability statement

The data that support the findings of this study are available in GenBank at https://www.ncbi.nlm.nih.gov/genbank/ under the Accession number OQ686796-OQ686799, OQ709803-OQ709858, and OQ709862-OQ709881. The clinical data not mentioned already in the manuscript is available from the corresponding author upon reasonable request.

## Ethics approval statement

This study was approved by the Institutional Ethics Committee of ICMR-National Institute of Cholera and Enteric Diseases, Kolkata.

