## Supplementary Table 1 for "Upsurge in Hospitalization of Pediatric Patients with Severe Acute Respiratory Infections in Kolkata and Surrounding Districts Caused by Recombinant Human Respiratory Adenovirus Type B 7/3"

**Supplementary Table 1**: **List of Primers used for amplification of Hexon and Fiber genes**

| Gene | Primer | Primer sequences | Position | Reference |
|---|---|---|---|---|
| Hexon | HAdV A F1 | 5'-CCTACTCTTACAAAGTTCG-3' | 18649-18667* | This study |
| | HAdV A R1 | 5'-CCCATATTTCCAGTACTGTT-3' | 19416-19435* | |
| | HAdV B F1 | 5'-TGGCCAGCACATTCTTTGACAT-3' | 18706-18727* | This study |
| | HAdV B R1 | 5'-TCAGTATTTCTGTCCTGCAA-3' | 19479-19498* | |
| Fiber | AdB1 | 5'-TSTACCCYTATGAAGATGAAAGC-3' | 31399-31419 | Xu et al., 2000 |
| | AdB2 | 5'-GGATAAGCTGTAGTRCTKGGCAT-3' | 32059-32079 | |

*Positions are in the reference genome of HAdV-B3 (GenBank acc. no. DQ099432)
